# Cooperative molecular mimicry drives prolonged autoinflammation in multisystem inflammatory syndrome in children

**DOI:** 10.64898/2026.04.03.26350001

**Authors:** Haley E Randolph, Ashley Richardson, Sofija Buta, Julie Samuels, Nina N Brodsky, Seunghee Kim-Schulze, Carrie L Lucas, Rebecca Trachtman, Dusan Bogunovic

## Abstract

Multisystem inflammatory syndrome in children (MIS-C) is a pediatric hyperinflammatory disease manifesting 4-6 weeks after SARS-CoV-2 infection. While the immunological hallmarks of MIS-C have been defined, few details regarding the underlying disease pathology have been resolved. To address this, we used a multiomics approach to profile the plasma and peripheral immune cells of 13 acute MIS-C patients, 18 recovered MIS-C follow-ups resampled over multiple time points (1-18 months), and 15 healthy pediatric controls. Despite rapid clinical disease resolution, circulating pro-inflammatory (IL-8, IL-6, IL-1α, IL-1β, TNF-β) and T_H_2-type cytokines (IL-4, IL-5, IL-13) remained elevated up to three months post-MIS-C onset, revealing a subclinical inflammatory state that endures in recovered children. Surprisingly, the majority of patient-expanded TCRs recognizing SARS-CoV-2 epitopes were cross-reactive (75%, 12/16 SARS-CoV-2 TCRs) for autoantigens related to prostaglandin biology and insulin metabolism, suggesting a breakdown of self-tolerance via SARS-CoV-2 molecular mimicry. Indeed, autoantibody screening confirmed that 13 gene targets with self-antigen peptides also exhibited elevated autoantibodies in MIS-C patients. Further, autoreactive TCR expansions lasted over time and correlated with cytokines involved in allergic inflammation. Together, our findings point to a mechanism of sustained autoimmunity wherein promiscuous TCRs recognize both viral and self-antigens that are activated during primary SARS-CoV-2 infection in children who develop MIS-C. Upon onset, these circulating cross-reactive T cells drive clinically apparent sterile autoinflammation that persists subclinically into convalescence.

## Main text

The consequences of SARS-CoV-2 infection in children are generally less severe compared to those seen in adults^1^. However, approximately four to six weeks after SARS-CoV-2 infection resolves, children can suddenly develop multisystem inflammatory syndrome in children (MIS-C), a rare but serious systemic pediatric hyperinflammatory disease characterized by acute inflammatory shock involving multiple organ systems, most notably the gastrointestinal, cardiovascular, hematologic, mucocutaneous, and respiratory systems^2^. Without prompt immunosuppressive treatment intervention, MIS-C can be life-threatening and rapidly progress to multiple organ failure and death^2,3^.

Similarities exist between MIS-C and other delayed hyperinflammatory syndromes, like Kawasaki disease, a rare pediatric vasculitis marked by high fever and rash^4^, and Kawasaki shock syndrome^5^, both of which are associated with prior viral exposure culminating in delayed systemic inflammation that very rarely recurs^6,7^. Although some symptomology is shared, acute MIS-C patients display distinct clinical and immune signatures. In particular, MIS-C cases present with increased circulating inflammatory cytokines and chemokines (particularly IL-6 and CXCL10)^8,9^ as well as a specific polyclonal expansion of activated T cells expressing the Vβ21.3 T cell receptor in a majority (> 60-70%) of patients^10–14^.

We and others have speculated that the etiology of MIS-C may involve an autoimmune component, given the delayed post-infection onset of the disease and the presence of certain autoantibodies in patients^8,9^. Recently, cross-reactive antibodies and T cells capable of engaging epitopes from both the SARS-CoV-2 nucleocapsid protein and SNX8, a protein critical to the regulation of the innate antiviral response^15^, were found in MIS-C patients^16^, suggesting that molecular mimicry induced by a viral trigger may play a role in disease progression. Further, it has been shown that upregulated serum TGF-β in MIS-C patients impairs cytotoxic Vβ21.3^+^ T cell responses and leads to Epstein-Barr virus (EBV) reactivation^11^, connecting MIS-C with underlying latent viral infection in a potentially causative manner.

To date, most studies investigating MIS-C have focused on characterizing molecular features of the intense, systemic inflammation seen in the primary response or mechanisms by which the Vβ21.3^+^ T cell expansion may give rise to hyperinflammation. Despite this, the definitive, causative pathobiology of MIS-C is still incompletely understood. In this study, we explore immune response dynamics in MIS-C, covering the period from acute onset to 18 months post-MIS-C, using a functional multiomics approach. Specifically, we examine how various immune signatures, including plasma cytokine levels, cellular markers of activation and exhaustion, global transcriptional patterns, paired T cell receptor repertoire dynamics, and circulating autoantibody levels, relate to one another and resolve in patients over time, demonstrating the advantages of longitudinal sampling in a clinical setting.

### Inflammatory and T_H_2-type cytokines remain elevated months after clinical resolution of MIS-C

In this study, we used paired single-cell RNA/T cell receptor (TCR)-sequencing together with high dimensional secreted cytokine (Luminex), mass cytometry (CyTOF), and/or autoantibody profiling to immunophenotype plasma and peripheral blood mononuclear cells (PBMCs) from a total of 13 MIS-C patients during the acute stage of disease, 18 samples obtained from recovered MIS-C patients resampled over a range of time points following initial MIS-C diagnosis (11 early follow-ups sampled at 1-3 months after MIS-C onset; 7 late follow-ups sampled at 6-18 months after MIS-C onset), and 15 healthy pediatric control donors (Fig. 1A, Table S1). Due to constraints on sample input material, not all assays were conducted on all samples (Table S1).

**Fig. 1.**
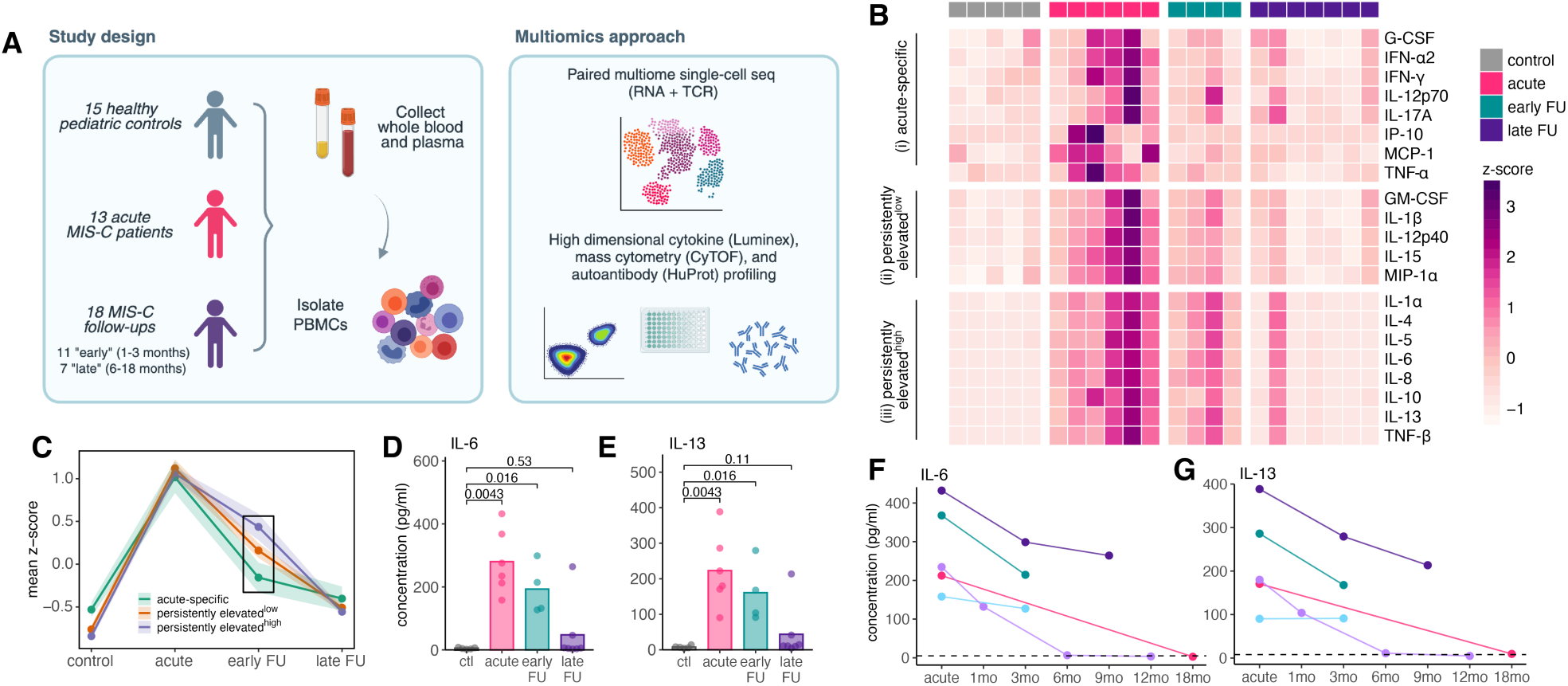
Cytokine signatures remain elevated up to 3 months after MIS-C onset. **(A)** Overview of the study design and multiomics approaches utilized to phenotype our MIS-C patient and follow-up cohort. **(B)** Heatmap of standardized cytokine concentrations across disease state groups (Luminex). Z-scores of cytokine concentrations were calculated across samples independently for each cytokine. Hierarchical clustering separates the cytokines into three classes: *(i)* acute-specific, *(ii)* persistently elevated^low^, and *(iii)* persistently elevated^high^. Only cytokines significantly elevated in acute patients compared to pediatric healthy controls are shown (p < 0.05, Mann-Whitney U tests). **(C)** Average trajectories for each of the cytokine classes determined in (A). Shaded area represents the mean ± standard deviation. **(D-E)** Representative persistently elevated^high^ cytokines, including **(D)** IL-6 and **(E)** IL-13. **(F-G)** Cytokine concentrations for **(F)** IL-6 and **(G)** IL-13 plotted by individual over time. Solid lines connect longitudinal samples from the same patient. Dashed lines represent the mean magnitude of each respective cytokine in the healthy pediatric controls. Mann-Whitney U tests were used to calculate the p-values in **(D-E)**.

While we and others have previously focused on delineating the primary effects of MIS-C at the systems-level, we now sought to understand how inflammatory signatures resolve in patients over time. To examine this, we assessed secreted cytokine profiles in acute MIS-C patients (n = 6) and follow-up samples taken at multiple time points over the recovery window (early follow-up: 1-3 months after MIS-C onset [n = 4]; late follow-up: 6-18 months after MIS-C onset [n = 7]) compared to healthy pediatric controls (n = 5). To measure the dynamic secreted immune response in MIS-C, we conducted high-dimensional plasma profiling for 30 cytokines and chemokines using the Luminex platform (Table S2).

Hierarchical clustering of the standardized cytokine concentrations across donors revealed three distinct classes of cytokines: *(i)* acute-specific, *(ii)* persistently elevated^low^, and *(iii)* persistently elevated^high^ (Fig. 1B). Acute-specific cytokines were elevated only in acute MIS-C patients (p < 0.05, control vs. acute), whereas persistently elevated^low^ and persistently elevated^high^ cytokines were significantly increased in both acute patient and early follow-up samples (p < 0.05 for both control vs. acute/early follow-up comparisons). Average trajectory analysis across cytokine clusters highlighted that the early follow-up time point alone was sufficient to distinguish the three different classes (Fig. 1C, Fig. S1A), with persistently elevated^high^ cytokines displaying the greatest magnitudes in early follow-up samples and acute-specific cytokines the lowest.

Persistently elevated^high^ cytokines were related to canonical pro-inflammatory processes (IL-6, IL-1α, IL-8, and TNF-β [Fig. 1B and D]) and T helper 2 (T_H_2) cell responses (IL-4, IL-5, and IL-13 [Fig. 1B and 1E]). Across patients, a sustained elevation of these markers was observed up to approximately three months post-MIS-C onset, with levels gradually returning to baseline afterwards (Fig. 1F-G). Persistently elevated^low^ cytokines also included pro-inflammatory cytokines as well as those involved in T cell activation and proliferation, such as IL-1β and IL-15 (Fig. 1B, Fig. S1B-C), which likewise returned to baseline levels after a period of heightened induction (Fig. 1B, Fig. S1D-E). These data point to a prolonged but subclinical sterile inflammation^8^ that lingers in children after recovering from acute disease, driven by dysregulated inflammatory and T_H_2-mediated cytokine responses known to be involved in autoimmunity^17–19^.

### Single-cell profiling reveals transcriptional remodeling of classical monocytes in MIS-C

We then evaluated the extent to which MIS-C influences the transcriptional response of immune cells over the convalescent period. To capture transcriptomic changes, we performed multiome single-cell RNA/TCR-sequencing on patient and follow-up PBMCs (n = 7 acute MIS-C patients, 5 early follow-ups, 7 late follow-ups, and 6 pediatric controls). To increase our acute and follow-up MIS-C cohort size, we integrated an independent, publicly available single-cell RNA/TCR-sequencing dataset consisting of 5 acute and 5 early follow-up MIS-C samples from Zhang et al.^12^ (Fig S2A, Table S1, detailed in Methods). Across individuals in the combined dataset, we profiled 260,356 single-cell transcriptomes (n = 40,290 cells from controls [n = 6], 78,320 cells from acute patients [n = 12], and 141,746 cells from follow-ups [n = 17]). Clustering followed by cell type label transfer annotation^20^ of the integrated dataset uncovered 30 different immune cell subsets (Fig. 2A).

**Fig. 2.**
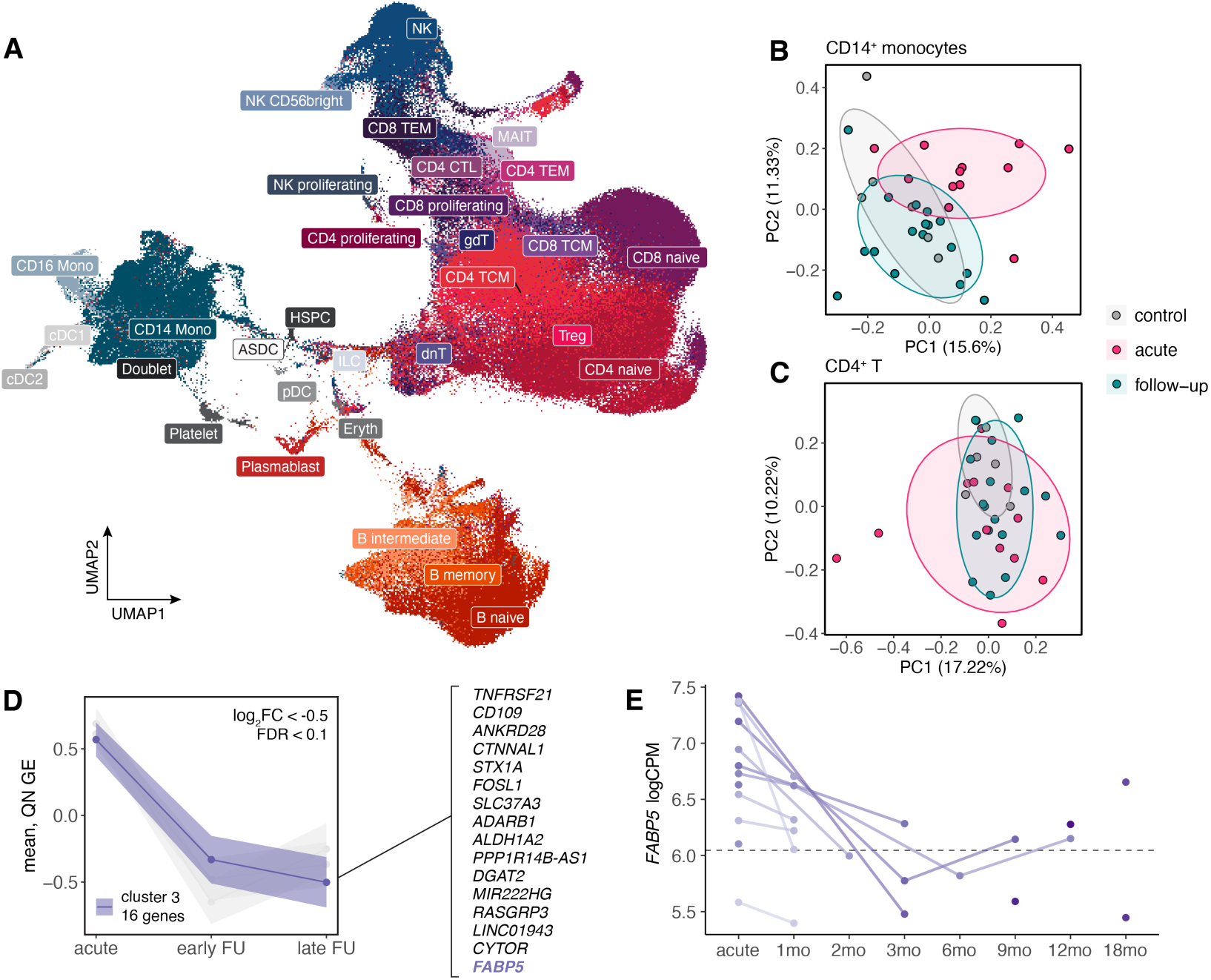
Transcriptional remodeling of CD14^+^ monocytes in acute MIS-C patients. **(A)** UMAP visualization of all cells (n = 260,356) profiled via scRNA-seq across healthy pediatric control, acute MIS-C patient, and follow-up samples. ASDC: AXL^+^SIGLEC6^+^ dendritic cells, CD4^+^ CTL: cytotoxic CD4^+^ T cells, cDC: conventional dendritic cells, dnT: double-negative T cells, Eryth: erythrocytes, gdT: gamma delta T cells, HSPC: hematopoietic stem and progenitor cells, ILC: innate lymphoid cells, MAIT: mucosal-associated invariant T cells, Mono: monocytes, NK: natural killer, pDC: plasmacytoid dendritic cells, TEM: T effector memory, TCM: T central memory. **(B-C)** PCA decompositions of the **(B)** CD14^+^ monocyte and **(C)** CD4^+^ T cell pseudobulk expression data colored by disease status. In **(B)** and **(C)**, expression estimates are corrected for the biological effects of age and sex and the technical effects of dataset, batch, and number of cells collected per sample. **(D)** Genes in CD14^+^ monocytes for which expression levels remain elevated in the early compared to late follow-ups (log_2_FC < −0.5, FDR < 0.1, cluster 3, hierarchical clustering on the quantile normalized expression values). Shaded area represents the mean ± 95% confidence interval. Gray lines represent the cluster 1 and cluster 2 trajectories. **(E)** Expression of *FABP5* across individuals. Samples from the same MIS-C patient over time are connected with colored lines. Dotted line represents the mean logCPM value for the healthy pediatric control group.

Next, we defined a broad set of immune cell populations corresponding to the major cell types found in PBMCs, including CD4^+^ T cells, CD8^+^ T cells, B cells, NK cells, CD14^+^ monocytes, and CD16^+^ monocytes. Within this set, we collapsed our single-cell gene expression estimates into pseudobulk estimates per sample. Principal component analysis (PCA) on these data revealed a clear separation of MIS-C cases from healthy pediatric control and convalescent follow-up samples only in CD14^+^ monocytes, reflected in the first principal component (PC) (percent variance explained [PVE] 15.6%) (Fig. 2B). All other cell types displayed a high degree of sample overlap across disease states (Fig. 2C, Fig. S2B-D). Notably, substantial transcriptional variation between MIS-C patients was observed across cell types (Fig. 2B-C; S2B-D), highlighting the large spectrum of clinical heterogeneity and disease severity previously described in MIS-C^8,21,22^.

To further tease apart dysregulated gene expression patterns in MIS-C, we modeled the effect of convalescence on expression, which allowed us to identify genes with expression levels linearly correlated with the dynamic transcriptional response, starting from acute disease and continuing throughout the recovery window (details in Methods). In line with our PCA analyses, CD14^+^ monocytes displayed the most obvious transcriptional changes (n = 303 response-associated genes, 3.0% of the transcriptome; |log_2_ fold change (FC)| > 0.5, false discovery rate [FDR] < 0.10), while the impact on other cell types was less evident (range: 0 −1.2% response-associated genes) (Fig. S2E, Table S3).

From the 303 response-associated genes in CD14^+^ monocytes, we focused on those upregulated in acute patients that gradually returned to baseline during the convalescent period (log_2_FC < −0.5, i.e., negative slope, n = 119 genes). Among these genes, we performed hierarchical clustering on the quantile normalized expression estimates to elucidate the qualitative shapes of their response trajectories without considering expression magnitude. Clustering revealed three distinct trajectories, one which contained a set of genes that remained elevated in early follow-ups (i.e., one to three months after MIS-C onset) compared to late follow-ups (n = 16 genes) (Fig. 2D), indicative of a prolonged, disease-associated transcriptional signature despite clinical resolution. This group contained several genes related to inflammation, immunometabolism, and cell signaling, including *FABP5* (Fig. 2E), which is involved in fatty acid transport and the regulation of inflammation through IL-1β/IL-17A signaling^23^ and the NF-κB pathway^24^. *CD109*, which encodes a TGF-β co-receptor and modulates both the TGF-β and NF-κB pathways^25^, also shares this trajectory. TGF-β overproduction has been implicated in the pathogenesis of MIS-C via impaired T cell cytotoxicity and subsequent EBV reactivation^11^. Together, these findings indicate that classical monocytes exhibit a long-lasting immunomodulatory gene signature that remains even after clinical disease has resolved.

### A loss of peripheral MAIT cells is a hallmark of MIS-C

Next, we sought to uncover perturbations in the T cell repertoire and define how TCR dynamics resolve over time. In our single-cell RNA/TCR-sequencing data, we captured a total of 131,149 T cells across all samples (Fig. S3A), of which paired αβ TCR sequence information was available for 102,247 T cells. We considered only those expressing a single α-chain (encoded by the *TRAV* genes) paired with a single β-chain (encoded by the *TRBV* genes), as these represent functional TCRs (n = 87,686 T cells). In addition, we restricted our analysis to common αβ TCR pairs found in ≥ 80% of the samples (n = 478 TCRs, 28 out of 35 samples) with the goal of probing shared receptors that shift proportions over time.

We first assessed paired αβ TCRs with decreased proportions in acute MIS-C patients compared to healthy pediatric controls (p < 0.05, Mann-Whitney U test) and detected ten TCR pairs significantly contracted in patients (Fig. 3A, Table S4). Of these ten receptor pairs, four carried a *TRAV1-2*-encoded α-chain paired with variable β-chains (Fig. 3A, bold), suggesting a coordinated reduction in *TRAV1-2*^+^ T cells. A subset of innate-like T cells known as mucosal-associated invariant T (MAIT) cells express this particular α-chain, and previous single-cell RNA-sequencing studies have used *TRAV1-2* expression as a marker of this population^26^. MAIT cells are characterized by a semi-invariant TCR α-chain comprised of *TRAV1-2* (Vα7.2) paired with a limited set of J-chain gene segments (most commonly *TRAJ33* [Jα33] but also *TRAJ20* [Jα20] and *TRAJ12* [Jα12])^27,28^. These non-MHC-restricted T cells play a role in immune surveillance and pathogen response by recognizing microbial vitamin B metabolites and can be activated by nonspecific inflammatory triggers^29,30^. They predominantly reside at mucosal surfaces, such as the gut lamina propria and lungs, but also circulate in the peripheral blood at lower proportions (1-10% of total T cells)^31,32^.

**Fig. 3.**
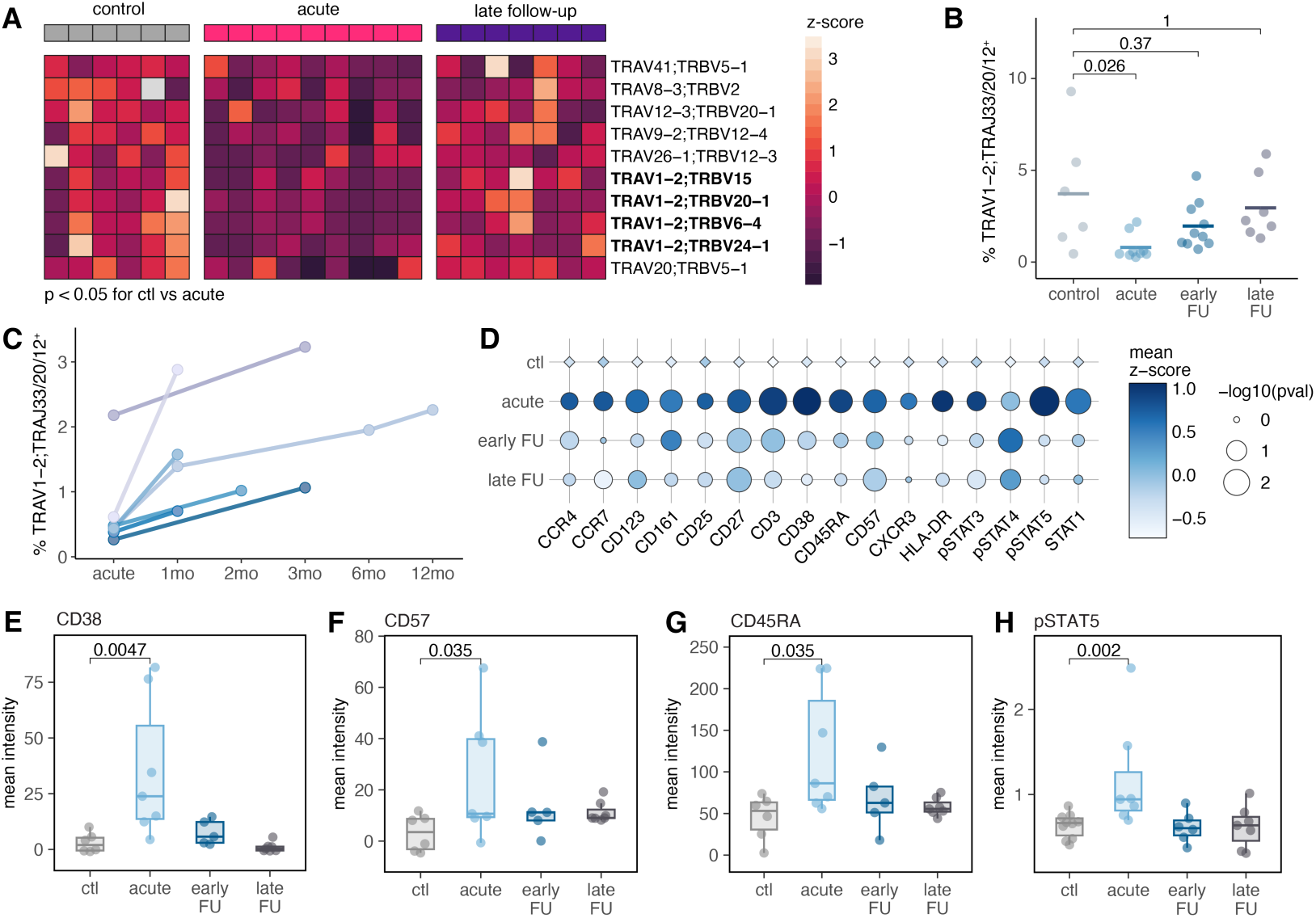
MAIT cells are contracted and functionally reprogrammed in acute MIS-C patients. **(A)** Heatmap of paired αβ TCRs that have significantly lower proportions in acute MIS-C patients compared to healthy pediatric controls (n = 10 TCRs, p < 0.05, Mann-Whitney U tests). Z-scores of TCR proportions were calculated across samples independently for each TCR. **(B)** Proportion of *TRAV1-2*;*TRAJ33*/*20*/*12*^+^ MAIT cells per sample stratified by sampling time point (scTCR-seq). **(C)** Proportion of *TRAV1-2*;*TRAJ33*/*20*/*12*^+^ MAIT cells over time for acute MIS-C patients with one or more follow-up time points (n = 7, scTCR-seq). Solid lines connect longitudinal samples from the same patient. **(D)** Bubble plot of the MDIPA and pSTAT panel markers (CyTOF) in MAIT cells. P-values are calculated on the mean fluorescence intensity (MFI) for samples in each disease state group against the healthy control group. Data are represented as the average standardized MFI per group. **(E-H)** Boxplots of various activation and exhaustion markers (CyTOF) in MAIT cells stratified by disease status. Mann-Whitney U tests were used to calculate the p-values in **(B)**, **(D)**, and **(E-H)**.

Because of the overall reduction of *TRAV1-2*^+^ T cells in patients, we more precisely pinpointed the MAIT cell population (defined as *TRAV1-2*;*TRAJ33*/*20*/*12*^+^ cells using our TCR-sequencing data) and asked how this population fluctuates over time. In line with our above analysis, we found decreased percentages of *TRAV1-2*;*TRAJ33*/*20*/*12*^+^ MAIT cells in MIS-C patients compared to healthy pediatric controls (p = 0.026) (Fig. 3B, Fig. S3B). Similarly, across patients with at least one follow-up time point, MAIT cell proportions increased as the duration from initial acute sampling increased (Fig. 3C), indicating that circulating MAIT cell frequencies decrease in the acute inflammatory state, potentially reflecting an egress of these cells from the periphery to sites of inflammation.

Next, we wanted to investigate functional changes in MAIT cells of patients, as phenotypic differences in these cells have been linked to numerous autoimmune, immune-mediated, and infectious disease contexts^33^. To do this, we performed mass cytometry profiling (CyTOF) with the Maxpar Direct Immune Profiling Assay (MDIPA)^34^, a 30-marker immunophenotyping panel, coupled with a phosphorylated STAT (pSTAT) panel to quantify intracellular JAK-STAT signaling (Table S5). MAIT cells were defined via surface markers (CD3^+^CD45^+^CD8^+^CD4^−^CD161^+^CD27^+^), and the fraction of MAIT cells per sample was highly concordant between our single-cell TCR-sequencing data and CyTOF data (Pearson *r* = 0.772, p = 6.6 x 10^-5^) (Fig. S3C).

Various markers of activation and exhaustion were upregulated in MAIT cells of MIS-C patients (Fig. 3D), including CD38 (11.7-fold increase over control, p = 0.0047) (Fig. 3E, Fig. S3D) and CD57 (7.9-fold increase over control, p = 0.035) (Fig. 3F, Fig. S3D). Increased surface expression of CD45RA was also observed on patient MAIT cells (p = 0.035) (Fig. 3G), which contradicts their typical effector memory phenotype^35^; however, populations enriched for naïve CD45RA^+^ MAIT cells have been reported in primary Sjögren’s syndrome (pSS), a chronic autoimmune disease^36^. Finally, intracellular pSTAT5 signaling (p = 0.002) (Fig. 3H) and STAT1 activity (p = 0.014) were elevated in patient cells, alluding to T cell activation and type I, II, or III interferon activation, which has previously been linked with functional impairment in MAIT cells^37^. Together, our results point to reduced peripheral circulation, functional immaturity, and a dual activation/exhaustion signature as common features of MAIT cells in acute MIS-C.

### Expanded TCRs that recognize SARS-CoV-2 epitopes are enriched for autoimmune signatures in MIS-C patients

Next, we identified paired αβ TCRs significantly increased in acute MIS-C patients compared to healthy pediatric controls, with the goal of detecting novel disease-associated T cell expansions. Other studies have underscored the importance of one particular polyclonal Vβ21.3 (encoded by the *TRBV11-2* gene) T cell expansion common to most (> 60-70%) MIS-C patients, which is characterized by a unique hyper-inflamed activation profile and linked with disease pathogenesis^10–14^. As validation, we first investigated the single-chain Vβ21.3^+^ expansion in our cohort and found enrichments in total CD8^+^ T cell (Fig. 4A) and CD8^+^ T effector memory (TEM) cell populations of patients (Fig. 4B). We then considered paired-chain TCR sequences and discovered 19 αβ TCR expansions in acute patients compared to pediatric control individuals (p < 0.05, Mann-Whitney U test) (Fig. 4C, Table S4). Four chains were among the most frequently represented in these expanded TCRs, including *TRBV11-2*, *TRAV14/DV4*, *TRAV29/DV5*, and *TRBV30* (n = 3 instances each), constituting 47.3% (9 out of 19) of all expanded αβ TCRs detected (Fig. 4C). Of note, the *TRBV11-2* chain was among these, suggesting that multiple polyclonal populations of Vβ21.3^+^ cells were expanded.

**Fig. 4.**
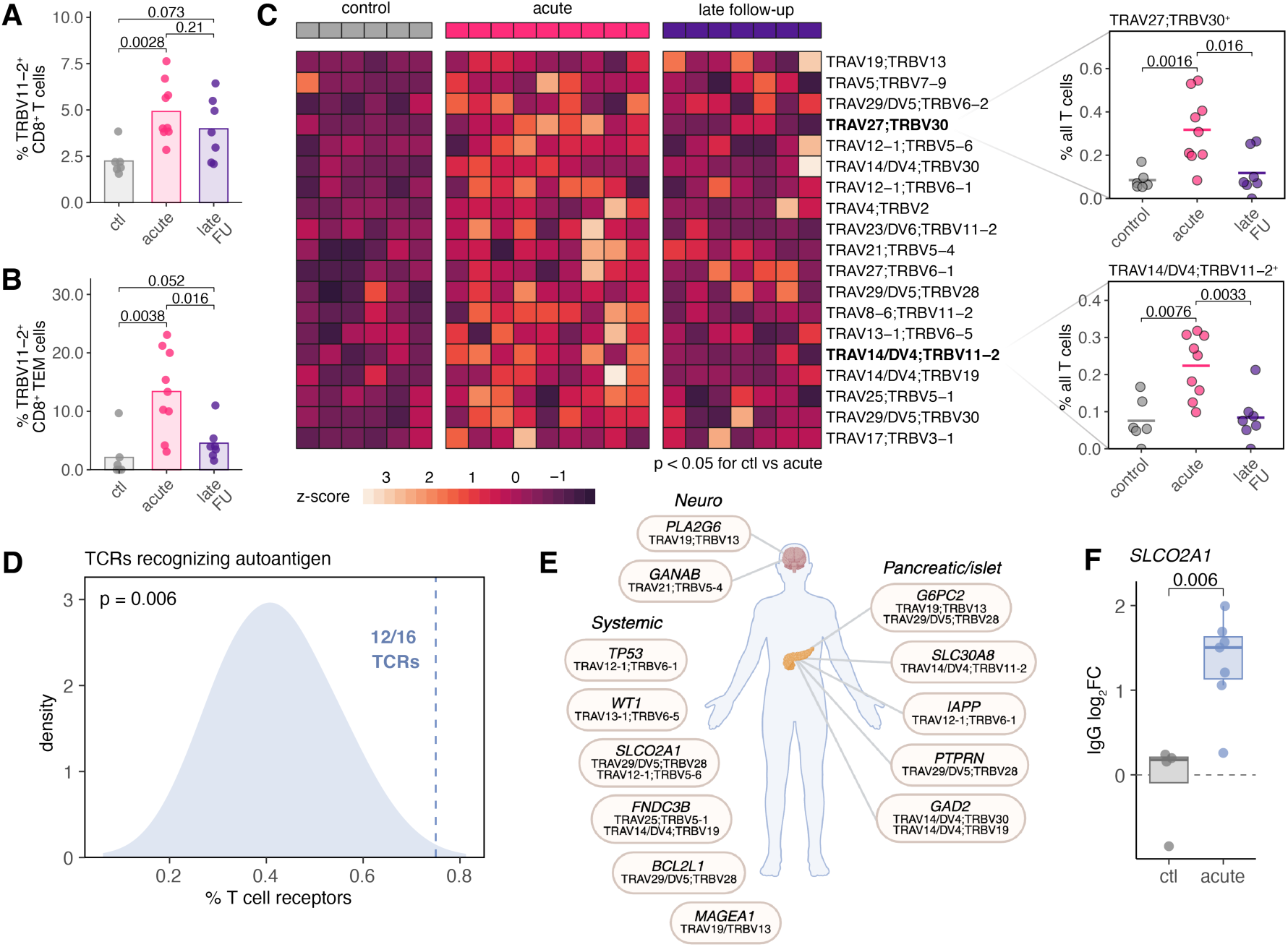
Expanded SARS-CoV-2-associated TCRs in acute MIS-C patients preferentially recognize autoantigens. **(A)** Proportion of *TRBV11-2*^+^ cells as the percentage of total CD8^+^ T cells by disease state group. **(B)** Proportion of *TRBV11-2*^+^ cells as the percentage of total CD8^+^ T effector memory cells by disease state group. **(C)** Heatmap of paired αβ TCRs that have significantly higher proportions in acute MIS-C patients compared to healthy pediatric controls (n = 19 TCRs, p < 0.05, Mann-Whitney U test). Z-scores of TCR proportions were calculated across samples independently for each TCR (left). Examples of expanded TCR pairs as a percentage of all T cells (right). **(D)** Observed proportion of MIS-C-expanded TCRs that recognize a SARS-CoV-2 epitope and at least one autoantigen epitope (12 out of 16, blue dotted line) compared to the null expectation when permuting random sets of TCRs that recognize SARS-CoV-2 epitopes of the same size (n = 16) and ascertaining how many also recognize at least one autoantigen epitope (n = 1,000 permutations, blue density distribution). **(E)** Schematic of the 13 autoimmune genes and their associated expanded TCR(s) with significantly elevated (p < 0.05) IgG autoantibodies in patients compared to healthy pediatric controls. **(F)** Example of one gene (*SLCO2A1*) with an autoimmune antigen that displays elevated IgG autoantibodies in MIS-C patients. Dotted line represents the y-intercept at zero. Mann-Whitney U tests were used to calculate the p-values in **(A-B)** and **(F)**.

Given the connection between cross-reactive T cells and MIS-C^16^, we hypothesized that a subset of these expanded TCRs may recognize self-antigens and represent autoreactive T cells. To test this, we annotated expanded TCR candidates using VDJdb^38^, a curated repository that couples known antigen specificities with paired-chain TCR sequences, by merging in annotations via defined *TRAV*;*TRBV* gene name pairs (Table S6). Of the expanded TCRs we identified in MIS-C patients, 84.2% (16 out of 19) recognized SARS-CoV-2 epitopes, indicative of expansions that were likely a direct consequence of the primary SARS-CoV-2 infection preceding MIS-C. Of the 16 TCRs recognizing SARS-CoV-2 epitopes, 75% (12 out of 16) also recognized at least one autoantigen, which is significantly higher than expected by chance (p = 0.006) as determined by a permutation approach (Fig. 4D, Methods).

To further investigate this autoimmune signature, we re-analyzed our previously reported data^8^ of plasma IgG and IgA reactivity in acute MIS-C (n = 7) and healthy pediatric (n = 4) donors against a microarray containing over 21,000 conformationally intact human peptides (HuProt Array). We focused our analyses on the 12 autoreactive TCRs identified above, which mapped to 17 different gene candidates with self-antigen peptides (Table S6). Of the 17 putative targets, MIS-C patients exhibited elevated IgG autoantibodies for 13 of these peptides compared to controls (Fig. 4E). These genes encoding self-antigens encompassed a wide range of functions, with their expression patterns distributed across the body and in tissues relevant to MIS-C pathology, such as the gastrointestinal tract, heart muscle, and lungs (Fig. 4E, Fig. S4A). One such gene, *SLCO2A1* (acute vs. control p = 6.0 x 10^-3^) (Fig. 4F), which is primarily expressed in the lungs, encodes a prostaglandin transporter responsible for moving prostaglandins and thromboxanes across cell membranes^39^. Interestingly, autoantibodies to *PTGS2*, encoding the enzyme COX-2 that plays a key role in the production of prostaglandins^40^, were also significantly elevated in the MIS-C group (acute vs. control p = 0.012) (Fig. S4B). Both prostaglandins and thromboxanes serve as inflammatory mediators that promote pain, fever, and inflammation and regulate blood clot formation, which is markedly disrupted in some MIS-C cases^41,42^. Additionally, *SLC30A8*, which encodes the ZnT8 zinc transporter involved in insulin secretion in pancreatic beta islet cells^43^, harbors self-peptides that triggered reactivity in our MIS-C cohort (acute vs. control p = 0.011) (Fig. S4C). Lastly, these patients displayed elevated autoantibodies to *SNX8* (acute vs. control p = 6.1 x 10^-3^) (Fig. S4D), which has recently been shown to drive viral molecular mimicry in the context of MIS-C^16^, and the SS-A/Ro 60kD antigen (acute vs. control p = 6.1 x 10^-3^), which is often elevated in individuals with systemic lupus erythematosus (SLE) and pSS^44^. Taken together, these data suggest that the expansion of SARS-CoV-2-specific but cross-reactive T cells may be a key contributor giving rise to systemic autoimmunity in MIS-C patients.

### Autoimmune-associated TCR expansions persist over time and correlate with cytokines involved in allergic inflammation

Finally, we hypothesized that circulating autoreactive T cell expansions may coincide with the prolonged inflammatory cytokine signatures that we observed earlier (Fig. 1B). To examine this, we probed the temporal dynamics of our patient-expanded cross-reactive T cell subset integrated with our cytokine profiling data (Table S2, Table S4). Among the autoreactive T cells expanded in MIS-C patients, four had proportions in acute patient and follow-up samples that were significantly associated (p < 0.05) with the magnitude of three or more cytokines, with *TRAV14/DV4*;*TRBV11-2^+^*and *TRAV29/DV5;TRBV28*^+^ T cells showing the most associations (Fig. 5A). Specifically, percentages of *TRAV14/DV4*;*TRBV11-2^+^* and *TRAV29/DV5;TRBV28*^+^ T cells were positively correlated with levels of the T_H_2-type cytokines IL-4, IL-5, and IL-13 and the pro-inflammatory cytokines IL-8, IL-6, IL-1α, IL-1β, and TNF-β (Fig. 5A, Fig. 5B). In addition to actively promoting inflammation, many of these cytokines are also connected to the pathogenesis of various autoimmune conditions, such as rheumatoid arthritis (RA), SLE, inflammatory bowel disease (IBD), and asthma, among others^17,18,45^. Interestingly, *TRAV14/DV4*;*TRBV11-2*^+^ T cells, which can recognize *SLC30A8* autoantigens (Fig. 4E), remained elevated in early follow-up samples compared to both healthy pediatric controls (p = 0.051) and late follow-up samples (p = 0.04) (Fig. S4E) and were significantly negatively correlated with months post-MIS-C onset (Pearson *r* = −0.478, p = 0.013) (Fig. 5C), revealing an association with disease resolution. Similarly, *TRAV29/DV5;TRBV28*^+^ cells, which can bind multiple self-antigens (*SLCO2A1*, *BCL2L1*, *G6PC2*, *PTPRN* [Fig. 4E]), also continued to be elevated in early follow-ups compared to pediatric controls (p = 0.011) (Fig. S4F). Notably, expanded autoreactive T cells were less anergic specifically in acute MIS-C patients compared to a background set of SARS-CoV-2-associated T cells (Fig. 5D), signifying an induced state of activation that arises only in the acute inflammatory environment. Together, this highlights the presence of cross-reactive, likely low-avidity T cell expansions associated with subclinical inflammation that continue into convalescence.

**Fig. 5.**
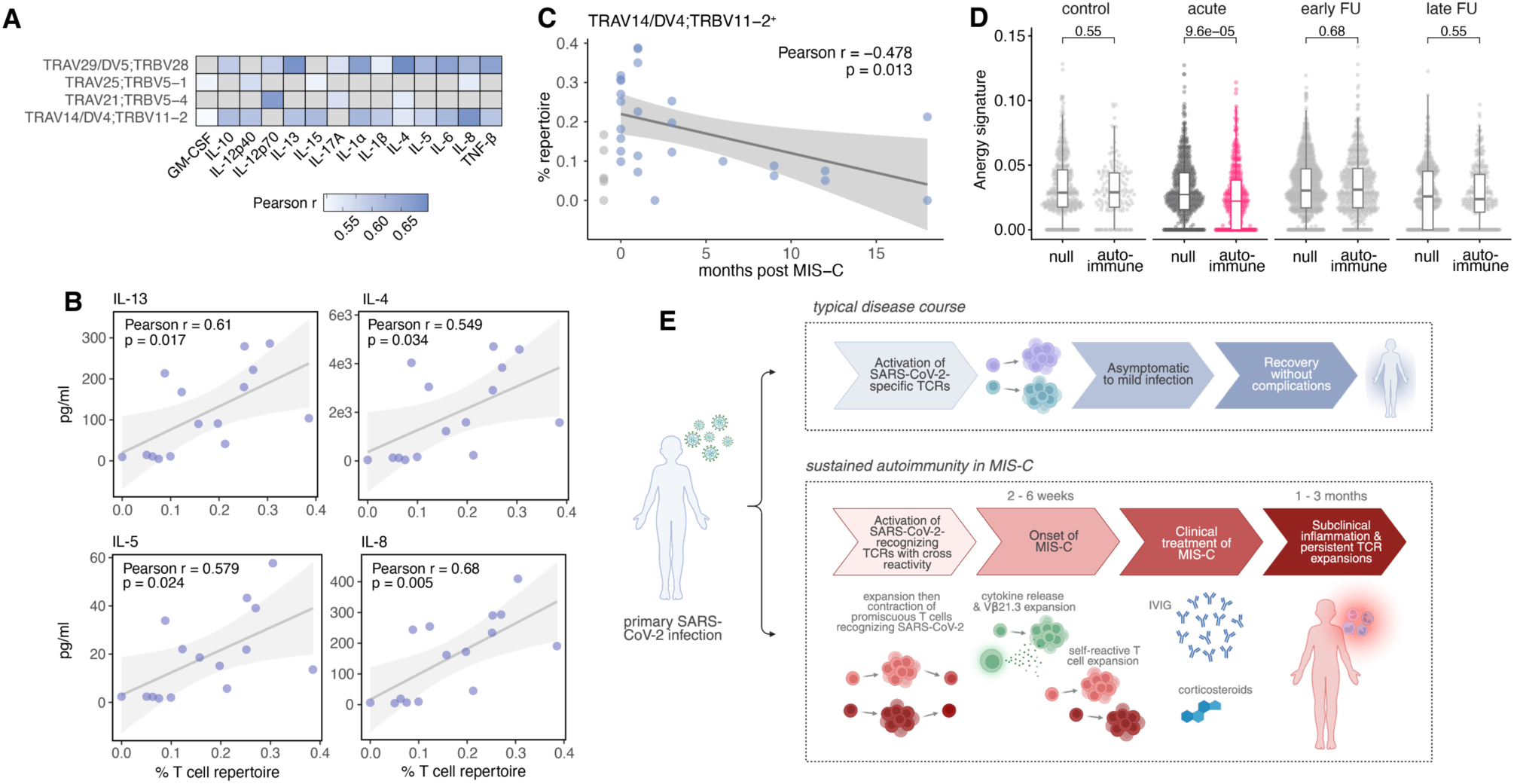
Autoimmune-associated TCR expansions persist months after disease resolution. **(A)** Pairwise comparison of the correlation (Pearson coefficient *r*) between the proportion of various expanded, autoreactive TCRs per sample and plasma cytokine levels (Luminex). Only TCRs with at least one significant (p < 0.05) association between proportion and cytokine concentration are shown. Only cytokines with greater than two significant (p < 0.05) associations across TCRs are shown. Correlations in gray are nonsignificant (p > 0.05). **(B)** The proportion of *TRAV14/DV4*;*TRBV11-2*^+^ T cells is correlated with inflammatory and T_H_2-type cytokines, including IL-13, IL-4, IL-5, and IL-8. **(C)** Correlation between months post-MIS-C and the proportion of *TRAV14/DV4*;*TRBV11-2*^+^ T cells (zero months corresponds to acute patients; healthy controls are shown in gray and are not included in linear regression). **(D)** Anergy module scores calculated from single-cell gene expression measurements for T cells with expanded autoimmune TCRs that also recognize SARS-CoV-2 (“autoimmune”, n = 12) compared to a random set that only recognize SARS-CoV-2 (“null”, n = 36) across time points (Mann-Whitney U tests). **(E)** Summary of the disseminated autoimmunity model in MIS-C. IVIG: intravenous immunoglobulin. In **(A)** and **(C)**, p-values and best-fit lines were obtained from linear regression models. The shaded area represents the mean ± 95% confidence interval.

## Discussion

Prior studies have implicated autoimmune mechanisms as one factor that may drive the pathology of MIS-C^8,9,16^. Our work supports this notion and builds on it, pointing to a circulating autoreactive T cell pool in children previously diagnosed with MIS-C. This persistent T cell repertoire coincides with sustained T_H_2 inflammatory responses in the subclinical, convalescent state well after the clinical resolution of acute inflammatory disease. We hypothesize that multiple polyclonal populations of SARS-CoV-2-recognizing T cells with self-cross-reactive TCRs expand during the primary SARS-CoV-2 infection in children who will go on to develop MIS-C, contrasting with the typical pediatric disease progression that generally involves an appropriate SARS-CoV-2-specific T cell response^46^ (Fig. 5E). Importantly in this model, primary infections for both the typical and pre-MIS-C outcomes are likely to be minor and without complication, agreeing with reports of asymptomatic to mild COVID-19 in children who do and do not develop MIS-C^8,47^; severe illness is likely only observed in the MIS-C group once MIS-C onsets.

Approximately four to six weeks later, these promiscuous, polyclonal self-reactive T cell populations expand and induce systemic autoimmune-driven pathological features that manifest as MIS-C, distinguished by heightened proinflammatory and T_H_2 cytokine release and Vβ21.3^+^ T cell expansions, among others (Fig. 4C, Fig. 5E). Why detrimental cross-reactivity may occur in few but not all children exposed to SARS-CoV-2 is not resolved here. We posit that the expansion of T cells prone to autoreactivity may be an aspect of most primary infections, but that progression to MIS-C likely depends on additional self and non-self determinants. These include the preexisting naïve T cell repertoire and MHC haplotypes expressed by an individual, the SARS-CoV-2 strain and specific viral peptide presented, and not yet fully defined environmental triggers, such as the presence of a secondary or latent viral infection, a co-occurring heightened inflammatory milieu, or a hapten^48^. Studies explicitly designed to investigate these possibilities are now needed. Because we detect multiple polyclonal expansions of putatively autoreactive T cells, it is tempting to speculate that, overall, this self-reactive T cell repertoire is what gives rise to the acute inflammation observed in MIS-C, implying that no clonal autoreactive T cell population alone is sufficient to induce disease. Indeed, this is in line with the multisystem presentation of MIS-C. If such a joint effect is necessary, this suggests that each single autoreactive T cell expresses a relatively low-avidity TCR, leading to a cooperative molecular mimicry.

One indication that this combinatorial pathogenesis conceivably plays a role comes from clinical observations. The first-line treatment for MIS-C remains high-dose corticosteroids, which are highly immunosuppressive in nature, paired with intravenous immunoglobulin (IVIG), which is also the treatment for related Kawasaki disease^49,50^. After administration, acute inflammation is rapidly quelled. The strong dose of steroids immediately blunts inflammatory processes and helps to resolve severe clinical manifestations, which may otherwise prove fatal, but is likely not sufficient to eliminate the autoreactive T cell pool. Although treatment alleviates the acute inflammatory assault, autoreactive T cell expansions persist, resulting in disseminated but diminished autoimmunity.

Additionally, we identified a decrease in the frequency of MAIT cells in the peripheral blood of acute MIS-C patients, which rebounded gradually over the convalescent period. This phenotype is not unique to MIS-C, but is also seen in other primary infectious diseases, such as SARS-CoV-2, influenza A virus, tuberculosis, and chronic HIV infection^51–55^, as well as various autoimmune conditions, including but not limited to RA, SLE, pSS, IBD, and multiple sclerosis^33,56^. This decline in frequency in the circulation likely signifies an egress from the periphery to target sites of inflammation, such as the airways^51^. In line with our findings, others have described a functional rewiring of MAIT cells in COVID-19, chronic HIV, and pSS patients that is marked by a simultaneous activation and exhaustion signature^36,52,55^. Of note, MAIT cells are canonically linked with mucosal immunity, and gastrointestinal involvement is a hallmark of MIS-C. Indeed, cytokines associated with barrier defense at the mucosa (IL-17A) and mucosal chemotaxis (CCL20 and CCL28) have been shown to be elevated in acute MIS-C patients^8^. Together, these data highlight an as yet unappreciated role of dysregulated MAIT cell responses in MIS-C, which is a recurring feature of certain other infectious and autoimmune diseases.

Our findings pertain not only to related childhood hyperinflammatory diseases, like Kawasaki disease, but also to other post-acute infection syndromes, such as long COVID, in which autoimmune components are suspected in at least a portion of cases^57^. Interestingly, parallels exist between these classes of disease, in particular the presence of sterile inflammatory sequalae and the presumption that molecular mimicry stemming from a latent or previous primary infection may play a role^16,57–59^. It will be important to delineate whether a syndrome like long COVID represents a form of MIS-C-like illness that presents in adults with significantly dampened peak inflammation that lingers over much longer time scales. Specifically, future studies will be needed to tease apart how autoimmune features, especially autoreactive T cell expansions, contribute to post-infectious syndromes and hyperinflammatory diseases, with the goal of uncovering potential shared disease etiologies.

## Data Availability

All raw sequencing data produced in the present study will be available upon formal journal publication. All other data produced in the present work are contained in the manuscript.

## Acknowledgements

We thank all study participants and the clinical research teams for their contributions. We thank Dr. Zhihong Chen, Geoffrey Kelly, and the team at the Human Immune Monitoring Center (HIMC) of the Icahn School of Medicine at Mount Sinai for their help with the 10x Genomics single-cell sequencing and CyTOF assays. This work was completed in part with Minerva computational resources provided by Scientific Computing and Data at the Icahn School of Medicine at Mount Sinai (Clinical and Translational Science Awards [CTSA] grant UL1TR004419, National Center for Advancing Translational Sciences). This work was also completed in part with computational resources provided by the Columbia University Center for Computational Biology and Bioinformatics (C2B2). Figures 1A, 4E, and 5E were created with BioRender.

## Author contributions

D.B. supervised the study. H.E.R. and D.B. designed the experiments. A.R., S.B., J.S., N.N.B, S.K., C.L.L., and R.T. provided patient samples and curated data. H.E.R. and A.R. performed experiments and sample collections. H.E.R. performed computational analyses. H.E.R. and D.B. wrote the manuscript, with input from all authors.

---

**Fig. S1.**
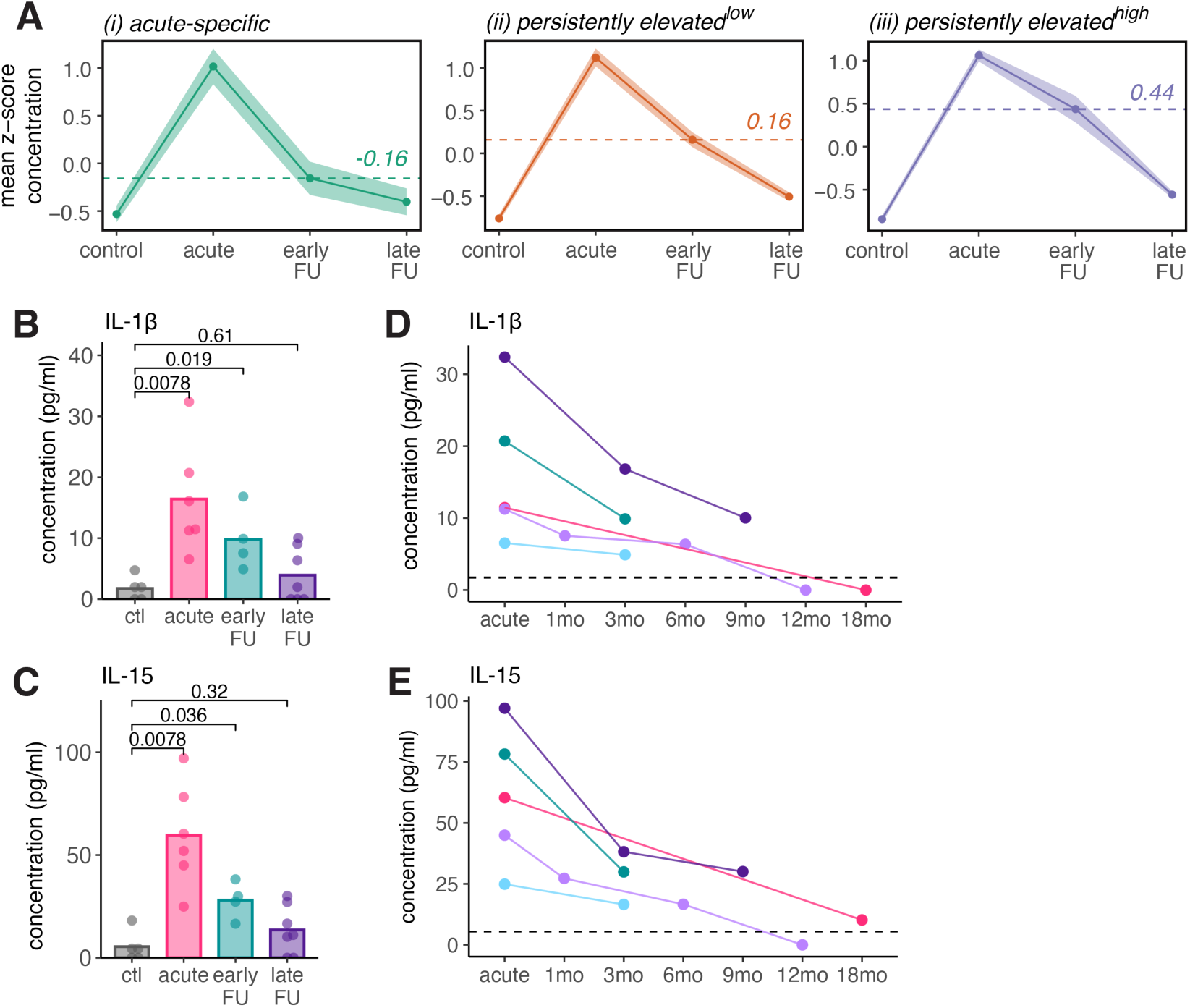
Cytokine signatures in MIS-C follow-ups. **(A)** Average trajectory for each of the cytokine classes determined in Fig. 1B split by cytokine class. Shaded area represents the mean ± standard deviation. Dashed lines represent the mean standardized concentration for the early follow-up group for each class. **(B-C)** Representative persistently elevated^low^ cytokines, including **(B)** IL-1β and **(C)** IL-15. **(D-E)** Cytokine concentrations for **(D)** IL-1β and **(E)** IL-15 plotted by individual over time. Solid lines connect longitudinal samples from the same patient. Dashed lines represent the mean magnitude of each respective cytokine in the healthy pediatric controls. Mann-Whitney U tests were used to calculate the p-values in **(B-C)**.

**Fig. S2.**
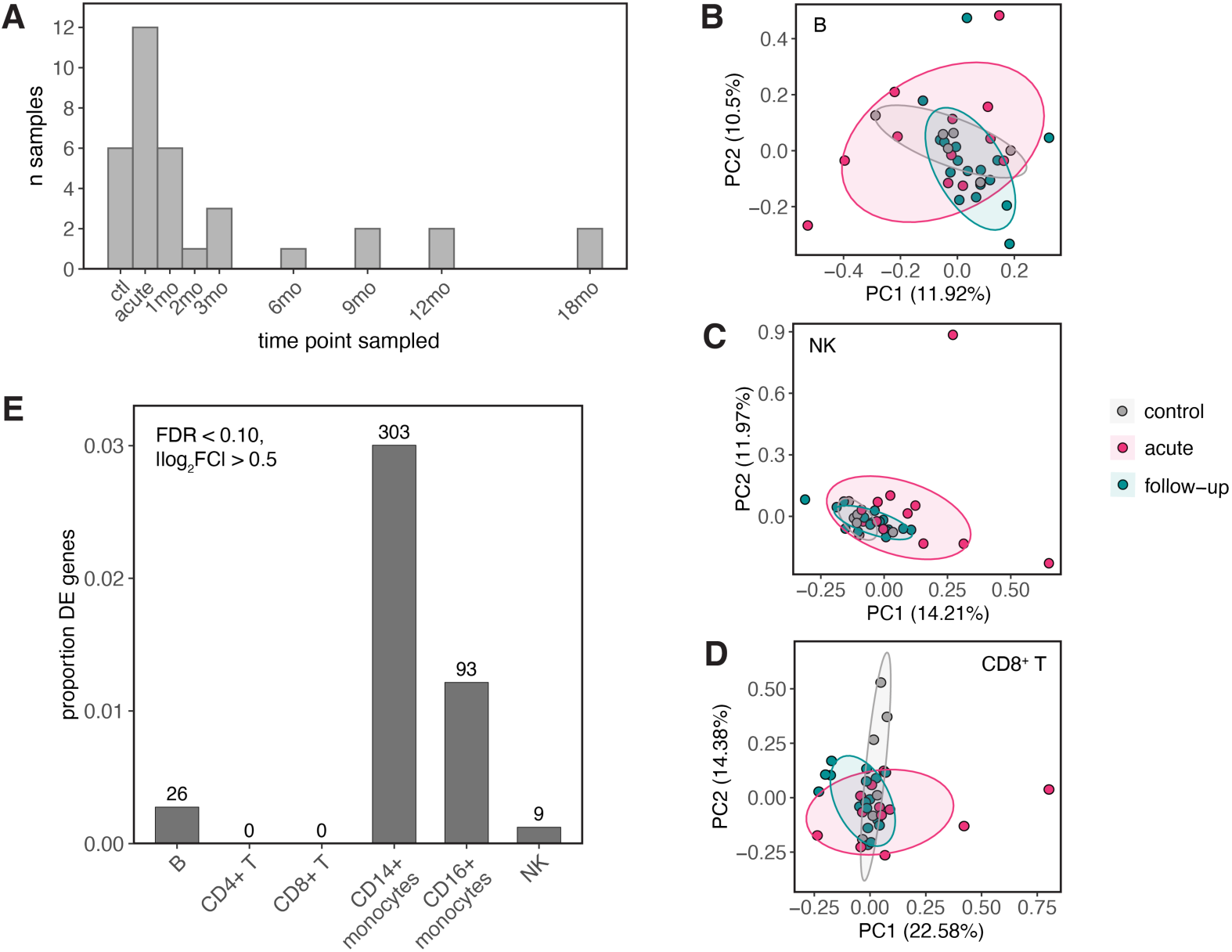
Sampling time points and global MIS-C effects on the transcriptome. **(A)** Distribution of sample collection time points across MIS-C patients in our single-cell multiome cohort. **(B-D)** PCA decompositions of the **(B)** B cell, **(C)** NK cell, and **(D)** CD8^+^ T cell expression data colored by disease status. In **(B-D)**, expression estimates are corrected for the biological effects of age and sex and the technical effects of dataset, batch, and number of cells collected per sample. **(E)** Numbers and proportions (y-axis) of significant MIS-C responsive genes (|log_2_FC| > 0.5, FDR < 0.10) across cell types.

**Fig. S3.**
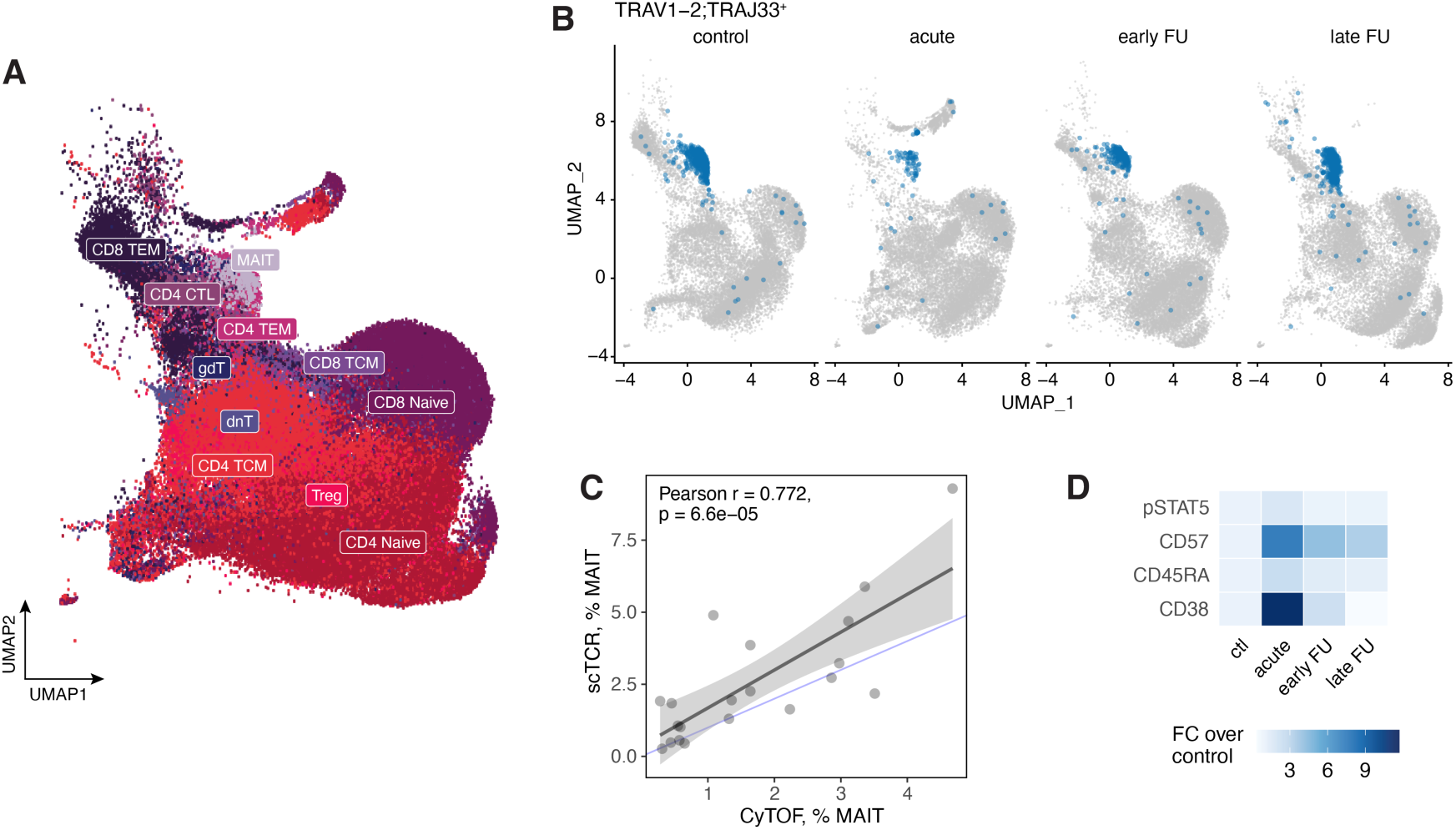
Changes in MAIT cell composition and function. **(A)** UMAP visualization of the T cells (n = 87,686) profiled via scRNA/TCR-seq across healthy pediatric control, acute MIS-C patient, and follow-up samples. CD4^+^ CTL: cytotoxic CD4^+^ T cells, dnT: double-negative T cells, gdT: gamma delta T cells, MAIT: mucosal-associated invariant T cells, TEM: T effector memory, TCM: T central memory. **(B)** UMAP visualization of *TRAV1-2*;*TRAJ33*^+^ MAIT cells, which are the most abundant J chain MAIT cell subtypes, stratified by disease state. Prior to visualization, within each disease state group, cells were randomly downsampled to the number of cells found in the least abundant group (healthy controls, n = 16,746 cells). **(C)** Correlation between the percent of MAIT cells identified via CyTOF (CD3^+^CD45^+^CD8^+^CD4^−^CD161^+^CD27^+^) versus scTCR-sequencing (*TRAV1-2*;*TRAJ33*/*20*/*12*^+^). P-value and best-fit line were obtained from a linear regression model. Shaded area represents the mean ± 95% confidence interval. Blue line represents the identity line. **(D)** Fold change of the average MFI values for MIS-C patients and follow-ups over healthy controls for selected CyTOF markers.

**Fig. S4.**
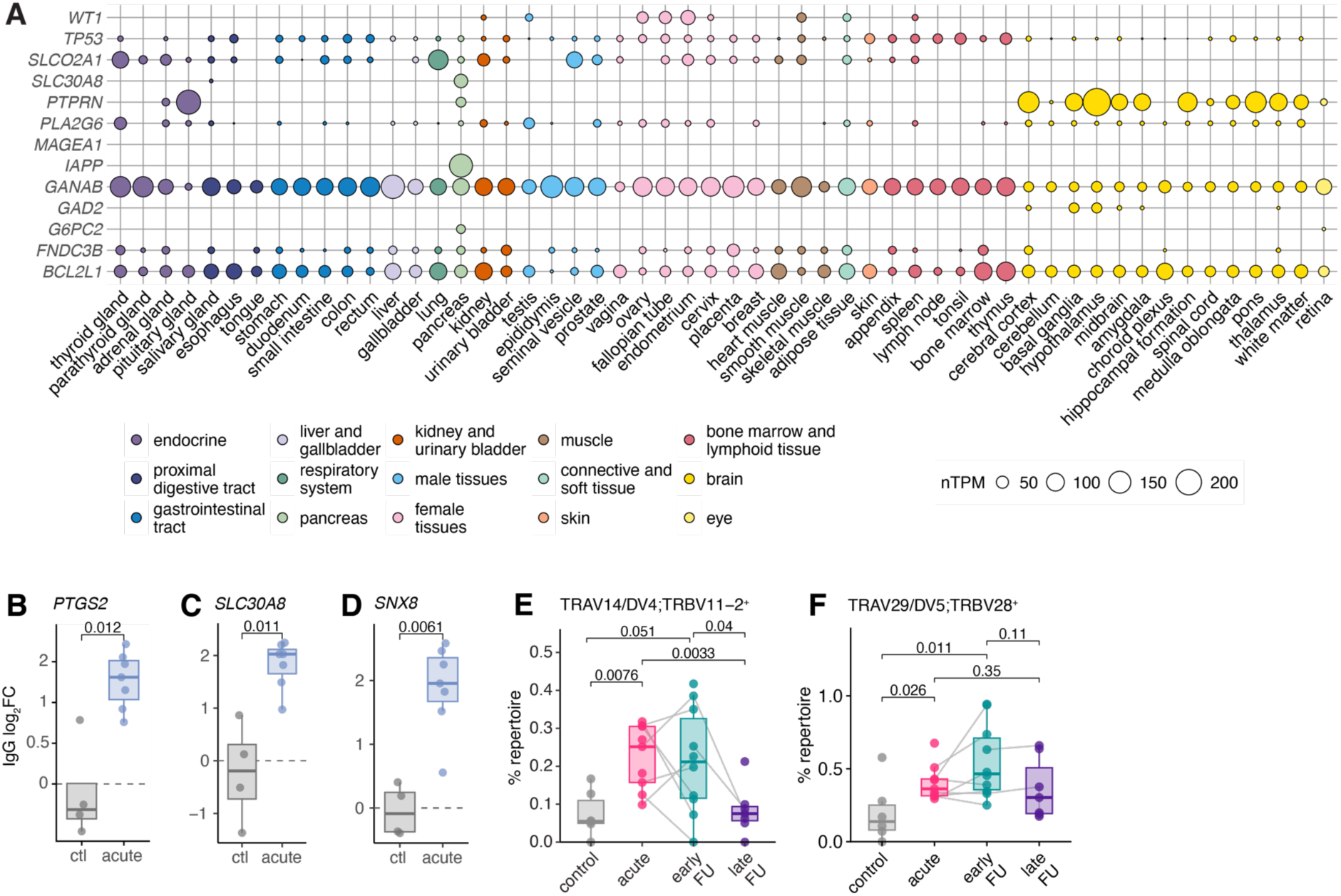
Autoimmune-associated gene, autoantibody, and TCR profiles. **(A)** Distribution of tissue-specific expression patterns for the autoimmune genes in which MIS-C patients display elevated IgG autoantibodies. Consensus transcript expression levels are summarized per gene in 54 tissues based on transcriptomics data from the Human Protein Atlas (HPA) and GTEx. **(B-D)** Examples of other autoimmune genes, including **(B)** *PTGS2*, **(C)** *SLC30A8*, and **(D)** *SNX8*, with elevated IgG autoantibodies in MIS-C patients. Dotted lines represent the y-intercepts at zero. **(E)** Percentage of *TRAV14/DV4*;*TRBV11-2*^+^ T cells per sample stratified by disease state group. **(F)** Percentage of *TRAV29/DV5*;*TRBV28*^+^ T cells per sample stratified by disease state group. In **(E-F)**, solid lines connect longitudinal samples from the same patient. Mann-Whitney U tests were used to calculate the p-values in **(B-F)**.

## Materials and Methods

### Participants and sample collections

Written informed consent for all participants was provided in compliance with Institutional Review Board protocols (Mount Sinai: STUDY-20-01314 and STUDY-20-01839, Columbia University: AAAV0619). All individual and sample IDs are de-identified and unknown to those outside of the research group. Acute MIS-C patients were recruited at the Mount Sinai Health System between April 1 through July 4, 2020, and convalescent samples were collected at various follow-up time points up to 18 months post-MIS-C onset. Blood was drawn into Cell Preparation Tubes (CPT) with sodium heparin (BD Vacutainer, cat no. 362753) and processed immediately. Peripheral blood mononuclear cells (PBMCs) and plasma were isolated via Histopaque-1077 separation (Sigma-Aldrich, cat no. H8889) and subsequently stored at −80C until use. PBMCs from healthy pediatric control donors living in New York City, USA were processed in parallel with MIS-C samples. Demographic data of recruited MIS-C patients and pediatric controls are detailed in Table S1. Clinical cohort data was previously published in Gruber et al. (2020)^8^. Healthy pediatric samples were age-matched to the extent possible (mean age_control_: 12.7 years, mean age_acute_: 9.7 years). Additionally, we computationally integrated a set of publicly available MIS-C patient and follow-up samples (n = 10) described in Zhang et al. (2024)^12^, which is detailed below (“Single-cell RNA-sequencing data processing”).

### Multiplex cytokine profiling via Luminex

Plasma cytokine measurements were assessed using the MILLIPLEX Human Cytokine/Chemokine Magnetic Bead 30-Plex Panel (MilliporeSigma, cat no. HCYTMAG-60K-PX30) according to manufacturer’s instructions. Undiluted supernatant samples were used as input, and analyte measurements were detected using the Luminex 200 Instrument. All samples were run on the same plate in duplicate, and duplicates were averaged per sample per analyte to obtain mean concentration values. Z-scores of the mean concentrations were calculated independently for each analyte across donors, and hierarchical clustering was performed on these standardized values to obtain the clustering trajectories shown in Figs. 1 and S1.

### Staining for mass cytometry profiling via CyTOF

Single-cell PBMC suspensions were barcoded using two panels: (1) the Maxpar Direct Immune Profiling Assay panel (MDIPA) (Standard Bio Tools, cat no. 201334) and (2) a custom phosphorylated STAT (pSTAT) panel (Table S5). Prior to staining, cell counts were recorded on the Revvity Cellaca MX Automated Cell Counter, and cell viability was measured using Acridine Orange/Propidium Iodide viability staining reagent (Revvity, cat no. CS2-0106).

For the MDIPA panel, cells were resuspended in Fc receptor blocking solution (BioLegend, cat no. 422302) and incubated for 10 min at room temperature. Fc blocked cells were then stained with the MDIPA panel following manufacturer’s instructions. For the pSTAT panel, Fc receptor blocking (BioLegend, cat no. 422302) and Rhodium-103 viability staining (Standard Bio Tools, cat no. 201103A) were performed simultaneously with surface marker staining for 30 min at room temperature. Cells were then washed twice in Cell Staining Buffer (CSB) (Standard Bio Tools, cat no. 201068), permeabilized with methanol, and stored at −80C overnight. The next day, cells were washed with CSB and blocked with heparin (100 units/mL) to prevent non-specific binding of intracellular antibodies^60^. Heparin-blocked cells were stained with phosphorylation markers on ice for 30 min. Separately for both panels after staining, cells were washed, fixed in 4.4% paraformaldehyde, and then labeled with 125 nM Iridium-193 (Standard Bio Tools, cat no. S00093) and 2 nM Osmium tetroxide.

### Acquisition and processing of CyTOF data in MAIT cells

Prior to data acquisition, samples were washed in Cell Acquisition Solution (Fluidigm, cat no. NC1919529) and resuspended at a concentration of 1 million cells/mL in Cell Acquisition Solution containing a 1:20 dilution of EQ Normalization Beads (Fluidigm, cat no. NC1307119). The samples were then acquired on a Helios mass cytometer equipped with a wide bore sample injector at an event rate of < 400 events per second. After acquisition, repeat acquisitions of the same sample were concatenated and normalized using Fluidigm software. FCS files were further cleaned using the Human Immune Monitoring Center (Mount Sinai) internal pipeline. Any aberrant acquisition time windows (three seconds) in which the cell sampling event rate was too high/low, defined as two standard deviations from the mean, were removed. Events from spike-in EQ Normalization Beads were removed, along with events that had low DNA signal intensity. Acquisition multiplets were also removed based on Gaussian parameters (“residual” and “offset”) acquired by the Helios mass cytometer.

After raw data quality filtering, FCS files were loaded into Cytobank^61^ and MAIT cells were gated as follows: CD3^+^CD45^+^CD8^+^CD4^−^CD161^+^CD27^+^. For functional markers in both the MDIPA and pSTAT panels, mean fluorescence intensity (MFI) values were extracted across samples. For each marker of interest, MFI values were corrected for experiment batch (two batches total) by fitting a model to estimate the effect of batch on MFI values (MFI ∼ 0 + batch) with the lm function in R (v4.0.3), taking the residuals of this model, and adding back the average batch effect (computed as the mean of the batch coefficients) to the residuals.

### Sample processing for single-cell sequencing

PBMCs were thawed in groups of five to eight samples, rested overnight (∼14 hours) in RPMI 1640 (Thermo Fisher, cat no. 21870092) supplemented with 10% FBS (Gibco, cat no. A52567-01), 1% GlutaMAX (Gibco, cat no. 35050-061), and 1% Penicillin-Streptomycin solution (Gibco, cat no. 15140-122), and subsequently processed for single-cell collection. For each sample, 10,000 cells were targeted for collection using the Chromium Single Cell 5’ Reagent Kit v2 Dual Index chemistry (10x Genomics, cat no. PN-1000244) paired with the Chromium Single Cell V(D)J TCR Amplification Kit (10x Genomics, cat no. PN-1000252). After droplet generation, the reverse transcription (RT) reaction was performed in a thermal cycler as described, and post-RT products were stored at −20C for up to one week until downstream processing. Post-RT reaction cleanup, cDNA/V(D)J amplification, and sequencing library preparation were performed as described in the Chromium Single Cell 5’ Reagent Kits v2 User Guide for both gene expression (GEX) and V(D)J modalities (10x Genomics, cat. no PN-1000190). Libraries were pooled and sequenced paired-end on an Illumina NovaSeq 6000 (median n cells captured per sample = 7,704, average median reads per cell = 29,338, average median genes detected per cell = 1,046).

### Single-cell RNA-sequencing data processing

Gene expression FASTQ files from each library were mapped to a prebuilt GRCh38 human reference (GRCh38-2020-A, downloaded from 10x Genomics) using cellranger (v7.0.1)^62^. Seurat (v4.3.0, R v4.0.3)^63^ was used to perform cell-level quality control filtering. High-quality cells were retained for downstream analysis if they had between 300 – 3500 genes detected (nFeature_RNA) and a mitochondrial UMI percentage < 12%, leaving 177,794 cells. Gene filtering was performed using the CreateSeuratObject min.cells parameter, in which only genes present in at least five cells were kept. To increase our sample size, we included a publicly available single-cell RNA/TCR dataset derived from acute and follow-up MIS-C samples (n = 10) described in Zhang et al., (2024).^12^ Similar cell-level filtering criteria were used for the Zhang et al. data (between 200 – 4000 nFeature_RNA and mitochondrial UMI < 12%), leaving 82,562 cells (n = 260,356 cells total across datasets). We then split the combined Seurat object by dataset and ran SCTransform^64^ to normalize and scale the UMI counts within dataset. We simultaneously regressed out variables corresponding to experiment batch, percent mitochondrial UMIs per cell, individual, and sampling time point in both datasets, and additionally, regressed out a cohort variable corresponding to the location in which a sample was obtained in our MIS-C data. We integrated both datasets together using the SelectIntegrationFeatures, PrepSCTIntegration, FindIntegrationAnchors, and IntegrateData framework^63^. After integration, dimensionality reduction was performed via UMAP (RunUMAP function, dims = 1:30) and PCA (RunPCA function, npcs = 30). A Shared Nearest Neighbor Graph was constructed using the FindNeighbors function (dims = 1:20, all other parameters set to default), and clusters were subsequently called using the FindClusters algorithm (resolution = 0.5, all other parameters set to default)^63^.

### Cell type assignment

We performed cell type annotation via label transfer to map cell type information onto our data. To perform the label transfer, we downloaded a multimodal human PBMC reference dataset as described in Hao et al.^20^. We followed the Seurat v4 Reference Mapping workflow, consisting of the FindTransferAnchors and MapQuery functions, with the Hao et al. reference dataset and the following parameters: normalization.method = “SCT” and reference.reduction = “spca”. Fine-scale populations derived from the predicted.celltype.l2 definitions were then collapsed into the following broad super populations encompassing the six major cell types found in PBMCs: CD4^+^ T cells = c(”CD4 CTL”, “CD4 Naive”, “CD4 Proliferating”, “CD4 TCM”, “CD4 TEM”, “Treg”), CD8^+^ T cells = c(”CD8 Naive”, “CD8 Proliferating”, “CD8 TCM”, “CD8 TEM”), NK cells = c(”NK”, “NK Proliferating”, “NK_CD56bright”), CD14^+^ monocytes = “CD14_monocytes”, CD16^+^ monocytes = “CD16_monocytes”, and B cells = c(”B intermediate”, “B memory”, “B naive”). In total, we annotated 260,356 high-quality cells across samples (n CD4^+^ T cells = 103,608, CD8^+^ T cells = 48,814, CD14^+^ monocytes = 28,629, CD16^+^ monocytes = 2,783, B cells = 42,645, NK cells = 18,811).

### Calculation of pseudobulk estimates

We summarized single-cell expression values into bulk-like expression estimates for downstream gene expression modeling. For each major cell type cluster (CD4^+^ T cells, CD8^+^ T cells, B cells, CD14^+^ monocytes, CD16^+^ monocytes, NK cells) per sample, raw UMI counts were summed across all cells assigned to that sample for each gene using the sparse_Sums function in textTinyR (v1.1.3)^65^, yielding an *n* x *m* expression matrix for each cluster, where *n* is the number of samples included in the study (*n* = 35) and *m* is the number of genes detected in the single-cell analysis (*m* = 29,279).

### Calculation of residuals for gene expression modeling

For each cell type, lowly-expressed genes were filtered using cell type-specific cutoffs (removed if median logCPM < 1.5 in CD4^+^ T cells and CD14^+^ monocytes, < 2.5 in B cells and CD8^+^ T cells, < 3.5 in CD16^+^ monocytes, and < 4.0 in NK cells), retaining the following number of genes: CD4^+^ T cells = 10,376, CD8^+^ T cells = 9,146, B cells = 9,448, CD14^+^ monocytes = 10,089, CD16^+^ monocytes = 7,651, and NK cells = 7,268. After removing lowly-expressed genes, normalization factors to scale the raw library sizes were calculated using calcNormFactors in edgeR (v 3.26.8)^66^. The voom function in limma (v3.40.6)^67^ was used to apply these size factors, estimate the mean-variance relationship, and convert raw pseudocounts to logCPM values. A model evaluating the technical effect of dataset (∼ 0 + dataset, where dataset corresponds to a factor variable defining samples in our dataset vs the Zhang et al. dataset) on gene expression was fit using the lmFit and eBayes functions, and model residuals were obtained using the residuals.MArrayLM function in limma^67^. The average dataset effect was computed by taking the mean of the dataset coefficients per gene and added back to the residuals across samples to generate dataset-corrected expression estimates. The voom inverse variance weights were included in the respective lmFit call for all models^67^.

### Modeling the effect of MIS-C recovery on gene expression

Only acute and follow-up MIS-C samples were retained for modeling the effect of MIS-C convalescence on gene expression (i.e., control samples were excluded). The following linear model was used to identify genes with expression levels linearly correlated with the recovery window of MIS-C:

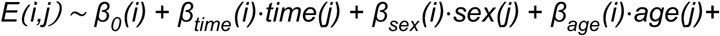

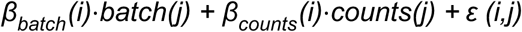

Here, *E(i,j)* represents the dataset-corrected expression estimate of gene *i* for individual *j*, *β_0_(i)* is the global intercept, and *β_time_(i)* represents the effect of the convalescent time on gene *i*’s expression. *Time(j)* represents the time period within the recovery window (treated as a numeric variable, where acute patients were coded as 0, early follow-ups as 1, and late follow-ups as 2). Age represents the mean-centered, scaled (mean = 0, sd = 1) age per individual, with *β_age_(i)* being the effect of age on expression levels; sex represents the self-identified sex for each individual, with *β_sex_(i)* capturing the effect of sex on expression; batch represents the experiment batch in which the sample was included, with *β_batch_(i)* capturing the batch effect; counts represents the number of cells captured within that cell type for sample *j*, with *β_counts_(i)* capturing the effect of cell number on expression; and *ε^cdt^* represents the residuals for each gene *i*, individual *j* pair. If age was missing for a patient (n = 1), it was filled with the average age across samples. The model was fit using the lmFit and eBayes functions in limma, and the estimates of *β_time_(i)* were extracted with their corresponding p-values. False discovery rates (FDR) were controlled using an approach analogous to that of Storey and Tibshirani^68^ using the empPvals and qvalue functions in the R package qvalue (v2.38.0)^69^, which derives the distribution of the null model empirically. To obtain a null, we performed ten permutations, where the *time(j)* label was permuted across individuals. We considered genes significantly associated with the convalescent period if |*β_time_*| > 0.5 and FDR < 0.10.

### Single-cell TCR-sequencing data processing

V(D)J FASTQ files from each library were mapped to a prebuilt GRCh38 human reference (vdj_GRCh38_alts_ensembl-5.0.0, downloaded from 10x Genomics) using cellranger (v7.0.1)^62^. Filtered contig annotation files were loaded into R (v4.0.3) using the repLoad function in “paired” mode from the R package immunarch (v0.10.3)^70^ The paired αβ chain data was converted to long format (i.e., each row was a unique T cell identified via cell barcode) for ease of subsequent processing. Prior to merging in TCR information, the Seurat object containing single-cell gene expression data was subset on only those T cell populations with > 500 cells total across samples (”CD4 CTL”, “CD4 Naive”, “CD4 TCM”, “CD4 TEM”, “CD8 Naive”, “CD8 TCM”, “CD8 TEM”, “Treg”, “MAIT”, “dnT”, “gdT”). TCR sequences were then merged into the Seurat object, and T cells missing chain annotations were removed (n = 30,334 cells). We also removed cells that did not visually cluster with the major T cell population on our UMAP (keep if UMAP1 > −4 & UMAP2 > −5), leaving 131,149 T cells with at least one detected α or β chain. We subsequently filtered our T cell dataset on the basis of the chain annotations themselves. First, we removed quadruple chain cells (ααββ: 1.3%) under the assumption that these were doublets. While we identified a large subset of T cells expressing only one chain (only α: 2.8% or only β: 19.2%), we subset these out because they likely do not represent functional T cells. We also removed T cells with triple chain annotations (ααβ: 6.1% or αββ: 2.5%), although the proportion is in line with previous human peripheral blood studies (∼10%)^71^. Finally, we removed three acute MIS-C samples with a relatively small number of annotated T cells (n < 900 cells per sample), giving us 87,686 T cells with paired αβ chain information to interrogate (median n paired αβ T cells across samples = 4111.5).

### Determination of TCR contractions and expansions

We identified a set of shared TCRs, designated as those present in at least 80% of the samples (n = 478 αβ TCRs), in which we subsequently probed variation in T cell repertoire dynamics. This criterion ensured that we examined relatively common TCRs found across samples (i.e., excluded very rare clonotypes) without requiring the stringent condition that a TCR be observed across all samples (n T cells expressing a shared TCR across samples = 53,444, 60.9%). Shared TCRs were defined by their unique *TRAV*;*TRBV* gene pair. For each sample independently, we calculated the proportion of each shared TCR in that sample as follows: *(i)* counted the number of *TRAV*;*TRBV*^+^ T cells observed in that sample for each shared TCR, and *(ii)* divided this count by the total number of shared TCR-expressing T cells for that sample. We defined a TCR as significantly contracted or expanded in acute MIS-C patients if the TCR proportion was significantly lower or higher in patients compared to healthy pediatric controls, respectively (p < 0.05, Mann-Whitney U tests).

### Annotation of expanded TCR sequences with validated TCR-epitope pairs

Paired αβ TCRs were annotated using VDJdb^38^, a database curated from publicly available literature that combines TCR sequence information with known antigen specificities. For each expanded TCR identified in Fig. 4C, we merged in VDJdb annotations by the defined *TRAV*;*TRBV* gene name pair. Afterwards, we considered whether expanded TCRs were annotated with recognized SARS-CoV-2 or autoantigen epitopes in the VDJdb Pathology column (respectively, “SARS-CoV-2” and “HomoSapiens”), and if annotated, which corresponding epitope gene was linked with that TCR (Fig. 4E).

### Computational enrichment of autoantibody signatures

To determine whether autoantigen epitopes were enriched among expanded, SARS-CoV-2-annotated TCRs, we used a permutation approach. In our data, the observed proportion of expanded, SARS-CoV-2-annotated TCRs that also recognized at least one autoantigen was 75% (12/16 TCRs tested). To empirically determine whether this proportion was more extreme than what would be expected by chance, we computed a null distribution from 1,000 permutations where, for each iteration, we: *(i)* sampled the same number of observed expanded, SARS-CoV-2-annotated TCRs (n = 16) from a background set of all SARS-CoV-2-annotated TCRs listed in VDJdb, *(ii)* obtained the number TCRs that also recognized at least one autoantigen from this background set, and *(iii)* calculated the null proportion of SARS-CoV-2- and autoantigen-annotated TCRs in this set. To compute a p-value, we evaluated the number of permutations in which the null proportion was greater than or equal to the observed proportion divided by the number of total permutations (n = 1,000).

### Calculation of anergy module score

To calculate anergy module scores for single T cells, we used the R package escape (v2.2.3)^72^, which integrates functions from the R package UCell (v2.10.1)^73^, a gene signature scoring method optimized for single-cell expression data. Anergy scores were calculated for two sets of T cells, the first being our set of expanded SARS-CoV-2-and autoimmune-annotated TCRs (n = 12, “autoimmune”) and the second being a random set of SARS-CoV-2-annotated TCRs defined by VDJdb (n = 36, “null”). The Seurat matrix was subset on the T cells in these defined sets, and the default Seurat assay was set to “SCT”. To compute module scores, we used the pre-defined anergy gene set available in escape (escape.gene.sets, name = “Anergy”, n = 31 genes) and the AddModuleScore_UCell function with default parameters.

## References

1. Brodin, P. SARS-CoV-2 infections in children: Understanding diverse outcomes. Immunity 55, 201–209 (2022).

2. Feldstein, L. R. et al. Multisystem Inflammatory Syndrome in U.S. Children and Adolescents. New England Journal of Medicine 383, 334–346 (2020).

3. Acevedo, L. et al. Mortality and clinical characteristics of multisystem inflammatory syndrome in children (MIS-C) associated with covid-19 in critically ill patients: an observational multicenter study (MISCO study). BMC Pediatr 21, 516 (2021).

4. Noval Rivas, M. & Arditi, M. Kawasaki disease: pathophysiology and insights from mouse models. Nat Rev Rheumatol 16, 391–405 (2020).

5. Gamez-Gonzalez, L. B. et al. Kawasaki disease shock syndrome: Unique and severe subtype of Kawasaki disease. Pediatr Int 60, 781–790 (2018).

6. Chang, L.-Y. et al. Viral infections associated with Kawasaki disease. Journal of the Formosan Medical Association 113, 148–154 (2014).

7. Goswami, N., Marzan, K., De Oliveira, E., Wagner-Lees, S. & Szmuszkovicz, J. Recurrent Kawasaki Disease: A Case Report of Three Separate Episodes at >4-Year Intervals. Children (Basel) 5, 155 (2018).

8. Gruber, C. N. et al. Mapping Systemic Inflammation and Antibody Responses in Multisystem Inflammatory Syndrome in Children (MIS-C). Cell 183, 982–995.e14 (2020).

9. Consiglio, C. R. et al. The Immunology of Multisystem Inflammatory Syndrome in Children with COVID-19. Cell 183, 968–981.e7 (2020).

10. Moreews, M. et al. Polyclonal expansion of TCR Vβ 21.3+ CD4+ and CD8+ T cells is a hallmark of multisystem inflammatory syndrome in children. Science Immunology 6, eabh1516 (2021).

11. Goetzke, C. C. et al. TGFβ links EBV to multisystem inflammatory syndrome in children. Nature 1–10 (2025) doi:10.1038/s41586-025-08697-6.

12. Zhang, Z. et al. Enhanced CD95 and interleukin 18 signalling accompany T cell receptor Vβ21.3+ activation in multi-inflammatory syndrome in children. Nat Commun 15, 4227 (2024).

13. Ramaswamy, A. et al. Immune dysregulation and autoreactivity correlate with disease severity in SARS-CoV-2-associated multisystem inflammatory syndrome in children. Immunity 54, 1083–1095.e7 (2021).

14. Porritt, R. A. et al. The autoimmune signature of hyperinflammatory multisystem inflammatory syndrome in children. J Clin Invest 131, (2021).

15. Guo, W. et al. SNX8 modulates the innate immune response to RNA viruses by regulating the aggregation of VISA. Cell Mol Immunol 17, 1126–1135 (2020).

16. Bodansky, A. et al. Molecular mimicry in multisystem inflammatory syndrome in children. Nature 632, 622–629 (2024).

17. Hirano, T. IL-6 in inflammation, autoimmunity and cancer. Int Immunol 33, 127–148 (2020).

18. Migliorini, P., Italiani, P., Pratesi, F., Puxeddu, I. & Boraschi, D. The IL-1 family cytokines and receptors in autoimmune diseases. Autoimmunity Reviews 19, 102617 (2020).

19. Singh, V. K., Mehrotra, S. & Agarwal, S. S. The paradigm of Th1 and Th2 cytokines: its relevance to autoimmunity and allergy. Immunol Res 20, 147–161 (1999).

20. Hao, Y. et al. Integrated analysis of multimodal single-cell data. Cell 184, 3573–3587.e29 (2021).

21. Diorio, C. et al. Proteomic profiling of MIS-C patients indicates heterogeneity relating to interferon gamma dysregulation and vascular endothelial dysfunction. Nat Commun 12, 7222 (2021).

22. Ma, K. C. et al. Phenotypic Classification of Multisystem Inflammatory Syndrome in Children Using Latent Class Analysis. JAMA Netw Open 8, e2456272 (2025).

23. Yu, J. et al. FABP5 in skin macrophages mediates saturated fat-induced IL-1β signaling in psoriatic inflammation. Cell Reports 44, 116254 (2025).

24. Vecchio, E. et al. FABP5 is a key player in metabolic modulation and NF-κB dependent inflammation driving pleural mesothelioma. Commun Biol 8, 324 (2025).

25. Batal, A., Garousi, S., Finnson, K. W. & Philip, A. CD109, a master regulator of inflammatory responses. Front Immunol 15, 1505008 (2025).

26. Shi, J. et al. Single-Cell Transcriptomic Profiling of MAIT Cells in Patients With COVID-19. Frontiers in Immunology 12, 700152 (2021).

27. Treiner, E. et al. Selection of evolutionarily conserved mucosal-associated invariant T cells by MR1. Nature 422, 164–169 (2003).

28. Reantragoon, R. et al. Antigen-loaded MR1 tetramers define T cell receptor heterogeneity in mucosal-associated invariant T cells. J Exp Med 210, 2305–2320 (2013).

29. Germain, L., Veloso, P., Lantz, O. & Legoux, F. MAIT cells: Conserved watchers on the wall. Journal of Experimental Medicine 222, e20232298 (2024).

30. Hinks, T. S. C. & Zhang, X.-W. MAIT Cell Activation and Functions. Front. Immunol. 11, (2020).

31. Martin, E. et al. Stepwise Development of MAIT Cells in Mouse and Human. PLOS Biology 7, e1000054 (2009).

32. Dusseaux, M. et al. Human MAIT cells are xenobiotic-resistant, tissue-targeted, CD161hi IL-17–secreting T cells. Blood 117, 1250–1259 (2011).

33. Toubal, A., Nel, I., Lotersztajn, S. & Lehuen, A. Mucosal-associated invariant T cells and disease. Nat Rev Immunol 19, 643–657 (2019).

34. Bagwell, C. B. et al. Multi-site reproducibility of a human immunophenotyping assay in whole blood and peripheral blood mononuclear cells preparations using CyTOF technology coupled with Maxpar Pathsetter, an automated data analysis system. Cytometry Part B: Clinical Cytometry 98, 146–160 (2020).

35. Nel, I., Bertrand, L., Toubal, A. & Lehuen, A. MAIT cells, guardians of skin and mucosa? Mucosal Immunol 14, 803–814 (2021).

36. Wang, J. J., Macardle, C., Weedon, H., Beroukas, D. & Banovic, T. Mucosal-associated invariant T cells are reduced and functionally immature in the peripheral blood of primary Sjögren’s syndrome patients. European Journal of Immunology 46, 2444–2453 (2016).

37. Tang, X. et al. Sustained IFN-I stimulation impairs MAIT cell responses to bacteria by inducing IL-10 during chronic HIV-1 infection. Science Advances 6, eaaz0374 (2020).

38. Shugay, M. et al. VDJdb: a curated database of T-cell receptor sequences with known antigen specificity. Nucleic Acids Research 46, D419–D427 (2018).

39. Xia, Z. et al. Structure and transport mechanism of the human prostaglandin transporter SLCO2A1. Nat Commun 16, 8124 (2025).

40. Simon, L. S. Role and regulation of cyclooxygenase-2 during inflammation. The American Journal of Medicine 106, 37S–42S (1999).

41. Hata, A. N. & Breyer, R. M. Pharmacology and signaling of prostaglandin receptors: Multiple roles in inflammation and immune modulation. Pharmacology & Therapeutics 103, 147–166 (2004).

42. Trapani, S. et al. Thromboembolic complications in children with COVID-19 and MIS-C: A narrative review. Front. Pediatr. 10, (2022).

43. Dwivedi, O. P. et al. Loss of ZnT8 function protects against diabetes by enhanced insulin secretion. Nat Genet 51, 1596–1606 (2019).

44. Yoshimi, R., Ueda, A., Ozato, K. & Ishigatsubo, Y. Clinical and Pathological Roles of Ro/SSA Autoantibody System. Clin Dev Immunol 2012, 606195 (2012).

45. Allard-Chamard, H. et al. Interleukin-15 in autoimmunity. Cytokine 136, 155258 (2020).

46. Hills, L. B., et al. Pediatric T cell and B cell responses to SARS-CoV-2 infection. JCI Insight 10, (2025).

47. Pierce, C. A. et al. Immune responses to SARS-CoV-2 infection in hospitalized pediatric and adult patients. Science Translational Medicine 12, eabd5487 (2020).

48. He, Z. et al. Food colorants metabolized by commensal bacteria promote colitis in mice with dysregulated expression of interleukin-23. Cell Metabolism 33, 1358–1371.e5 (2021).

49. McArdle, A. J. et al. Treatment of Multisystem Inflammatory Syndrome in Children. New England Journal of Medicine 385, 11–22 (2021).

50. McCrindle, B. W. et al. Diagnosis, Treatment, and Long-Term Management of Kawasaki Disease: A Scientific Statement for Health Professionals From the American Heart Association. Circulation 135, e927–e999 (2017).

51. Parrot, T., et al. MAIT cell activation and dynamics associated with COVID-19 disease severity. Sci Immunol 5, eabe1670 (2020).

52. Flament, H. et al. Outcome of SARS-CoV-2 infection is linked to MAIT cell activation and cytotoxicity. Nat Immunol 22, 322–335 (2021).

53. van Wilgenburg, B. et al. MAIT cells are activated during human viral infections. Nat Commun 7, 11653 (2016).

54. Jiang, J. et al. Mucosal-associated invariant T-cell function is modulated by programmed death-1 signaling in patients with active tuberculosis. Am J Respir Crit Care Med 190, 329–339 (2014).

55. Leeansyah, E. et al. Activation, exhaustion, and persistent decline of the antimicrobial MR1-restricted MAIT-cell population in chronic HIV-1 infection. Blood 121, 1124–1135 (2013).

56. Fan, Q. et al. New insights into MAIT cells in autoimmune diseases. Biomedicine & Pharmacotherapy 159, 114250 (2023).

57. Talwar, S., Harker, J. A., Openshaw, P. J. M. & Thwaites, R. S. Autoimmunity in long COVID. Journal of Allergy and Clinical Immunology 155, 1082–1094 (2025).

58. Marrani, E., Burns, J. C. & Cimaz, R. How Should We Classify Kawasaki Disease? Front. Immunol. 9, (2018).

59. Klein, J. et al. Distinguishing features of long COVID identified through immune profiling. Nature 623, 139–148 (2023).

60. Rahman, A. H., Tordesillas, L. & Berin, M. C. Heparin reduces nonspecific eosinophil staining artifacts in mass cytometry experiments. Cytometry A 89, 601–607 (2016).

61. Kotecha, N., Krutzik, P. O. & Irish, J. M. Web-based analysis and publication of flow cytometry experiments. Curr Protoc Cytom Chapter 10, Unit10.17 (2010).

62. Zheng, G. X. Y. et al. Massively parallel digital transcriptional profiling of single cells. Nat Commun 8, 14049 (2017).

63. Stuart, T. et al. Comprehensive Integration of Single-Cell Data. Cell 177, 1888–1902.e21 (2019).

64. Hafemeister, C. & Satija, R. Normalization and variance stabilization of single-cell RNA-seq data using regularized negative binomial regression. Genome Biology 20, 296 (2019).

65. Mouselimis, L. textTinyR: Text Processing for Small or Big Data Files. (2023).

66. Robinson, M. D., McCarthy, D. J. & Smyth, G. K. edgeR: a Bioconductor package for differential expression analysis of digital gene expression data. Bioinformatics 26, 139–140 (2010).

67. Ritchie, M. E. et al. limma powers differential expression analyses for RNA-sequencing and microarray studies. Nucleic Acids Res 43, e47–e47 (2015).

68. Storey, J. D. & Tibshirani, R. Statistical significance for genomewide studies. PNAS 100, 9440–9445 (2003).

69. Storey, J. D., Bass, A. J., Dabney, A., Robinson, D. & Warnes, G. qvalue: Q-value estimation for false discovery rate control. Bioconductor version: Release (3.16) 10.18129/B9.bioc.qvalue (2023).

70. Popov, A., et al. immunarch: An R Package for Painless Bioinformatics Analysis of T-Cell and B-Cell Immune Repertoires. ImmunoMind Team 10.5281/zenodo.10840553 (2024).

71. Zhu, L. et al. scRNA-seq revealed the special TCR β & α V(D)J allelic inclusion rearrangement and the high proportion dual (or more) TCR-expressing cells. Cell Death Dis 14, 1–11 (2023).

72. Borcherding, N. et al. Mapping the immune environment in clear cell renal carcinoma by single-cell genomics. Commun Biol 4, 122 (2021).

73. Andreatta, M. & Carmona, S. J. UCell: Robust and scalable single-cell gene signature scoring. Comput Struct Biotechnol J 19, 3796–3798 (2021).

